# Going off the rails: Impaired coherence in the speech of patients with semantic control deficits

**DOI:** 10.1101/2020.02.10.20020685

**Authors:** Paul Hoffman, Lucy Cogdell-Brooke, Hannah E Thompson

**Affiliations:** School of Philosophy, Psychology & Language Sciences, University of Edinburgh, UK; School of Psychology, University of Surrey, UK

**Keywords:** speech, discourse, aphasia, semantic control, coherence

## Abstract

The ability to speak coherently, maintaining focus on the topic at hand, is critical for effective communication and is commonly impaired following brain damage. Recent data suggests that executive processes that regulate access to semantic knowledge (i.e., semantic control) are critical for maintaining coherence during speech. To test this hypothesis, we assessed speech coherence in a case-series of stroke patients who exhibited deficits in semantic control. Patients were asked to speak about a series of topics and their responses were analysed using computational linguistic methods to derive measures of their global coherence (the degree to which they spoke about the topic given) and local coherence (the degree to which they maintained a topic from one utterance to the next). Compared with age-matched controls, patients showed severe impairments to global coherence but not to local coherence. Global coherence was strongly correlated with the patients’ performance on tests of semantic control, with greater semantic control deficits associated with poorer ability to maintain global coherence. Other aspects of speech production were also impaired but were not significantly correlated with semantic control deficits. These results suggest that semantic control deficits give rise to speech that is poorly regulated at the macrolinguistic “message” level. The preservation of local coherence in the patients suggests that automatic activation of semantic associations is relatively intact, such that each utterance they produce is connected meaningfully to the next. However, in the absence of control processes to constrain semantic activation, the content of their speech becomes increasingly distant from the original topic of discourse. This study is the first to investigate the impact of semantic control impairments on speech production at the discourse level and suggests that patients with these impairments are likely to have difficulties maintaining coherence in conversation.

## Introduction

Engaging in discourse is a complex cognitive activity, in which a speaker must identify the topic under discussion, generate a series of statements relevant to this subject and monitor their speech as the discourse unfolds to ensure that they remain on-topic. Discourse that successfully navigates these challenges is said to be coherent: it consists of a series of well-connected statements all related to a shared topic, making it easy to comprehend (Foltz, 2007; Glosser & Deser, 1992). Researchers often distinguish between global coherence, the degree to which each utterance relates to the overall topic under discussion, and local coherence, the degree to which adjoining utterances related meaningfully to one another (Glosser & Deser, 1992; Kintsch & Vandijk, 1978). Speech that frequently goes “off the rails”, deviating greatly from the original topic, is said to be low in global coherence, while speech that that lacks meaningful links from one utterance to the next is low in local coherence.

The ability to speak coherently is frequently impaired following brain damage, even in patients who do not display classical language production deficits (Ellis et al., 2016). Deficits to global coherence have been reported following stroke (Barker et al., 2017; Christiansen, 1995; Karaduman et al., 2017; Rogalski et al., 2010; Wright & Capilouto, 2012). Poor coherence is also a characteristic of speech in Alzheimer’s disease (Glosser & Deser, 1990), traumatic brain injury (TBI) (Glosser & Deser, 1990; Marini et al., 2014) and the frontal variant of frontotemporal dementia (Ash et al., 2006). In addition, healthy ageing is associated with declines in global and local coherence. Older adults are more likely to produce tangential, off-topic utterances in conversation (Arbuckle & Gold, 1993; Glosser & Deser, 1992) and to provide irrelevant information when telling a story (Juncos-Rabadan et al., 2005; Marini et al., 2005) or describing an object (Long et al., 2018). These impairments in coherence hinder effective communication and can be associated with reduced well-being and decreased satisfaction with social interactions (Arbuckle & Gold, 1993; Gold et al., 1988; Pushkar et al., 2000).

Coherence is a feature of the macrolinguistic structure of language – the way in which concepts and messages are organised within a whole discourse (Glosser & Deser, 1990; Marini et al., 2011). This macro level is often contrasted with the microlinguistic level, which is concerned with the phonological, lexical and grammatical characteristics of individual utterances. Coherence is often impaired in aphasic patients who have prominent microlinguistic impairments. Andreetta and Marini (2015) studied the narrative speech of 20 patients with fluent aphasia and found increased rates of global and local coherence errors, relative to healthy controls. Similar results have been reported in other studies of people with aphasia and it has been suggested that speech in these patients lacks coherence because their microlinguistic errors disrupt the flow of speech and the ability to construct a coherent narrative (Andreetta et al., 2012; Christiansen, 1995; Wright & Capilouto, 2012). On the other hand, some studies of aphasic patients have found normal levels of coherence in their speech production (Glosser & Deser, 1990; Ulatowska et al., 1983) and, conversely, impaired coherence has frequently been observed in the speech of non-aphasic people without microlinguistic deficits (Barker et al., 2017; Coelho, 2002; Glosser & Deser, 1990; Marini et al., 2014). Therefore poor coherence is not solely a consequence of disruption to other levels of the language system.

What other cognitive impairments could give rise to impaired coherence in speech? Research has mainly focused on the role of domain-general cognitive and executive systems, rather than language-specific processes, in the planning and regulation of speech content (Alexander, 2006; Barker et al., 2020; Barker et al., 2017; Kintz et al., 2016). In TBI patients, for example, poor coherence is observed in the context of minimal impairments to microlinguistic aspects of speech, such as phonology, lexical and syntactic processing (Coelho, 2002; Marini et al., 2014). In these patients, however, correlations have been reported between performance on the Wisconsin card-sorting test and errors of local and global coherence (Marini et al., 2014) and more general impairments of narrative organisation (Coelho, 2002), suggesting that an executive deficit may underpin their difficulties in regulating their speech. Similarly, coherence deficits in frontotemporal dementia are present in patients with executive impairment but not in those with the non-fluent or semantic variants of the disorder (Ash et al., 2006). In healthy individuals, age-related coherence declines have also been attributed to impaired executive function (Gold et al., 1988; Kintz et al., 2016; North et al., 1986).

These results suggest that domain-general executive control processes are involved in the monitoring and selection of topics during speech. One view holds that poor coherence, at the global level in particular, results from a reduced ability to inhibit irrelevant information, such that people are less able to prevent irrelevant or off-topic ideas from becoming automatically activated and intruding into their discourse (Arbuckle & Gold, 1993; Marini & Andreetta, 2016). Recent work has developed this hypothesis by investigating the specific role of *semantic control* in maintaining coherence. Semantic control refers to control processes that govern the retrieval and selection of semantic knowledge (Hoffman, McClelland, et al., 2018; Jefferies, 2013; Lambon Ralph et al., 2017). Much research has focused on how semantic control operates at the level of single words. For example, neuroimaging studies report similar left frontotemporal increases in activation when participants perform comprehension tasks that require them to inhibit semantic information that is irrelevant to the current context (Badre et al., 2005), to select the contextually-appropriate meaning of a homonym (Vitello & Rodd, 2015), and to resolve competition when multiple words compete for selection in naming tasks (Thompson-Schill et al., 1997). Deficits in these abilities have all been observed in patients suffering from multimodal semantic impairments following stroke (Jefferies et al., 2007; Jefferies & Lambon Ralph, 2006; Noonan et al., 2010).

The role of semantic control has rarely been studied in the context of discourse-level speech production. In recent work, however, Hoffman et al. (2018) hypothesised that semantic control processes are critically involved in maintaining coherence because semantic knowledge is central to all propositional speech. This is obviously true at the lexical level, since the selection of words for production is determined by their meanings. But at the broader “message” level, the content of our speech is also determined by our general semantic knowledge about the topic under discussion. For example, if someone asks you how they would catch a train somewhere, you must access stored semantic knowledge about the typical characteristics of railway stations, the methods for buying tickets and so on. To ensure coherent discourse, this knowledge retrieval process must be regulated such that topic-relevant knowledge is used to drive speech production and any irrelevant associations that come to mind are avoided (e.g., retrieving information about trains might bring to mind other forms of transport not relevant to the discussion). For these reasons, we predicted that the coherence of an individual’s discourse is influenced by the efficiency of their semantic control processes, in addition to more general executive function.

Hoffman et al. (2018) tested these predictors in a group of 60 healthy young and older adults. In line with previous studies, we found that the coherence of speakers was predicted by their performance on a test of domain-general executive function (the Trails test). Importantly, however, we also found that semantic control abilities were a significant independent predictor of coherence. Specifically, participants who produced more coherent speech performed better on a semantic decision task that required them to inhibit irrelevant semantic knowledge. These results suggest overlap in the semantic control processes operating at the lexical level and the macrolinguistic message level.

The results of Hoffman et al. (2018) suggest that semantic control processes, particularly those governing the selection of task-relevant semantic knowledge, contribute to the maintenance of coherence in healthy individuals. In the present study, we investigated whether this was also true for neuropsychological patients experiencing semantic control deficits. We investigated coherence in a case-series of patients presenting with multimodal semantic deficits following left-hemisphere stroke. Previous research has shown that semantic impairments in these individuals are typically a consequence of poor semantic control (Jefferies & Lambon Ralph, 2006; Lambon Ralph et al., 2017; Noonan et al., 2010). In the first part of the study, we established that this was also the case in our group. We then went on to collect samples of patients speaking about different topics and determined their coherence using previously-validated methods from computational linguistics (Hoffman, Loginova, et al., 2018). We predicted that patients would display impaired coherence, relative to healthy controls, and that the severity of semantic control impairment would predict the degree to which coherence was disrupted in individual patients. By assessing discourse-level speech in patients with semantic control deficits for the first time, we also aimed to establish how these deficits impact on patients’ extended speech production.

In addition to our main focus on semantic impairments following left-hemisphere stroke, we tested one individual who suffered a right-hemisphere stroke. Deficits in global coherence have often been reported following right-hemisphere stroke and some researchers have suggested that macrolinguistic processes critical for coherence are more dependent on the right hemisphere (Barker et al., 2017; Bartels-Tobin & Hinckley, 2005; Davis et al., 1997; Karaduman et al., 2017; Sherratt & Bryan, 2012). We therefore hypothesized that the right-hemisphere patient might experience impairments in coherence. As this patient presented with no semantic control deficits, if they showed a coherence deficit this would suggest that there are alternative mechanisms by which coherence can be impaired.

## Method

### Participants

Seven chronic left-hemisphere stroke patients were recruited from Yorkshire, Surrey, Sussex and Manchester community stroke groups.^1^ All suffered a left-hemisphere stroke at least four years previous to the study. In line with the inclusion criteria adopted by Jefferies and Lambon Ralph (2006), we recruited patients who showed difficulties accessing semantic knowledge in both verbal and non-verbal tasks (details given below). Previous work has shown that when multimodal semantic deficits occur following left-hemisphere stroke, these are typically a consequence of impaired semantic control (Jefferies & Lambon Ralph, 2006; Lambon Ralph et al., 2017; Noonan et al., 2010). Therefore we expected that this group would have difficulty controlling their access to semantic knowledge and we confirmed this in the first part of the study. In keeping with previous work, we refer to these participants as patients with semantic aphasia (SA) as, in addition to multimodal semantic deficits, they tend to present with other language impairments. Because we selected patients purely on the basis of their multimodal semantic impairments, we did not obtain formal diagnoses of aphasia or classifications of aphasia type. However, previous studies using the same inclusion criteria as ours have found that SA patients present with a range of different aphasia types (Jefferies & Lambon Ralph, 2006; Thompson et al., 2018).

MRI scans were available for five of the seven SA patients (see Table 1 for description of lesion locations and Supplementary Figure 1 for scan images). Patients presented with damage to a range of left-hemisphere sites. This is in line with previous studies showing that similar semantic control deficits can arise from either left prefrontal or posterior temporal and parietal damage (Jefferies & Lambon Ralph, 2006; Noonan et al., 2010; Thompson et al., 2018).

**Table 1:**
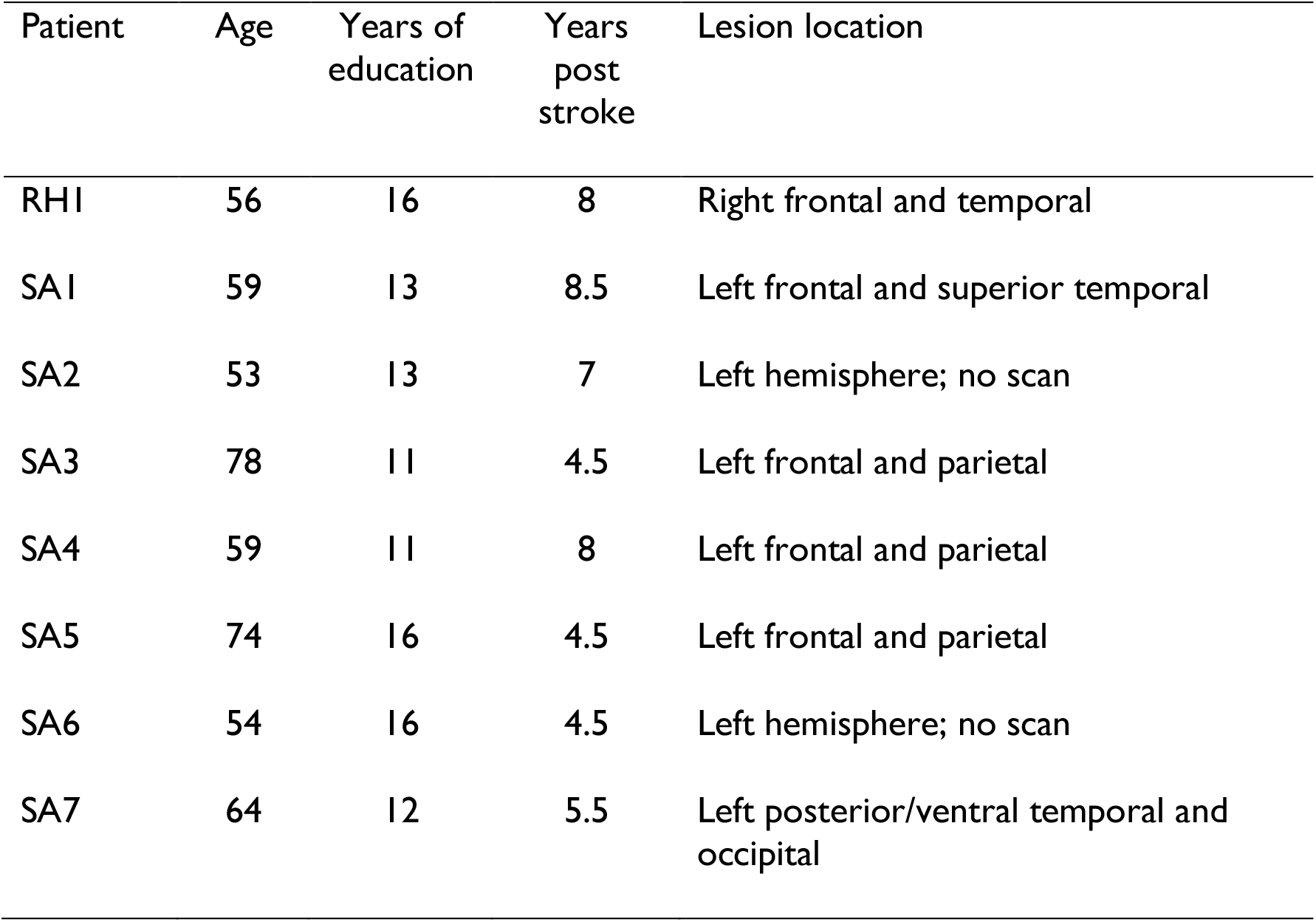
Patient demographic and lesion information

We also included one right-hemisphere stroke patient as a control case. This individual had suffered a right-hemisphere stroke but did not show the semantic deficits observed in the SA patients.

Age, years of education and time since stroke for all patients are reported in Table 1. All patients provided informed written consent to participate in this study. Ethical approval was obtained from the University of Surrey Ethics Committee (UEC/2016/090/FHMS). It also received ethical approval from the NRES Committee Yorkshire and The Humber (ref: 12/YH/0323). No patients were receiving speech and language therapy at the time of the study.

The characteristics of speech in the patients were compared with control data from healthy older adults, taken from Hoffman et al. (2018). The control sample comprised all participants reported by Hoffman et al. (2018) who were aged between 60 and 75 (N=12). The control group were matched to the SA patients for age (SA mean = 63.0 years; control mean = 67.4 years; *t*(17) = 1.44, *p* = 0.17) and years of education (SA mean = 13.1 years; control mean = 13.5 years; *t*(17) = 0.31, *p* = 0.76).

### Background neuropsychological tests

To be included in the study, SA patients had to perform below the normal range of healthy individuals in at least one verbal and one non-verbal test of semantic processing. To test verbal semantic processing, we used a 96-item synonym judgement task (Jefferies et al., 2009). In this comprehension task, patients were presented with a word and chose the word most similar in meaning from three options (e.g., is FROG similar to JEWEL, PICKLE or TOAD?). Two non-verbal semantic tests were conducted. In the Camel and Cactus Test (Bozeat et al., 2000), patients were presented with a picture and four options, one of which was semantically associated with it (e.g., does CAMEL go with CACTUS, TREE, FLOWER or ROSE?). The Object Use Test (Corbett et al., 2011) involved selecting an object for a task (e.g., BASH A NAIL INTO WOOD). The target was a picture of either the canonical tool (e.g., HAMMER), or a non-canonical object that could be used (e.g., BRICK), presented among a set of five unrelated distractors.

Other tests of language processing included two components of the Cambridge 64-item semantic test battery (Bozeat et al., 2000): spoken word–picture matching using 10 semantically-related response options, picture naming The Cookie Theft picture description test was run (Goodglass, 1983), giving a measure of propositional speech rate for each patient. Additionally, category and letter fluency tasks were run, using eight categories (animals, fruit, birds, breeds of dog, household objects, tools, vehicles, types of boat) and three letters (F, A and S).

Patients were also examined on a range of general neuropsychological assessments, including forwards and backwards digit span (Wechsler, 1987), dot counting and number location from the Visual Object and Space Perception (VOSP) battery (Warrington & James, 1991), the Coloured Progressive Matrices test of non-verbal reasoning (Raven, 1962), The Trail Making Test part A and B (Reitan, 1992) and the Brixton Test of Spatial Anticipation (Burgess & Shallice, 1997).

### Tests of semantic control

Semantic control was assessed using a 2 × 2 manipulation of semantic control demands in two different tasks (Hoffman, 2018) (following Badre et al., 2005). In the first task, participants made semantic decisions based on global semantic association. They were presented with a probe word and asked to select its closest semantic associate from either two or four alternatives (see Supplementary Figure 2 for examples). The strength of relationship between the probe and target was manipulated: the associate was either strongly associated with the probe (e.g., *town-city*) or weakly associated (e.g., *iron-ring*). The Weak Association condition was assumed to place greater demands on semantic control, specifically the *controlled retrieval* of semantic information, as automatic spreading of activation in the semantic network would be insufficient to identify the correct response (Badre & Wagner, 2007).

In the second task, participants were asked to match items based on specific features. At the beginning of each block, participants were given a feature on which to base their decisions (e.g., colour). On each trial, they were given the name of an object and were asked to select another item that was most similar on the specified feature. We manipulated the semantic congruency of the probe and target. On Congruent trials, the probe and target shared a pre-existing semantic relationship, as well matching on the currently relevant feature (e.g., *cloud-snow* are semantically related in addition to matching in colour). In contrast, on Incongruent trials the probe and target shared no meaningful relationship, other than their similarity on the specified feature (e.g., *salt-dove* are both typically white but otherwise semantically unrelated). Moreover, on Incongruent trials one of the foils had a strong semantic relationship with the probe, but did not match on the currently relevant feature (*salt-pepper*). Incongruent trials placed high demands on semantic control, and particularly on *semantic selection* processes, for two reasons. First, because there was no pre-existing semantic relationship between probe and target, participants could only identify the correct response if they focused selectively on the pre-specified features of the items and not on their other semantic properties. Second, participants were required to inhibit the strong but irrelevant relationship between the probe and foil.

### Speech elicitation

Procedures for obtaining speech samples was based on those used by Hoffman et al. (2018) with healthy participants, but were adapted for use with people with aphasia. Hoffman et al. presented participants with 14 different prompts to elicit speech on different topics. To avoid patient fatigue, we used only six prompts here:

1. Describe the steps you would need to take if going somewhere by train.
2. What do the police do when a crime has been committed?
3. Which is your favourite season and why?
4. Do you think it’s a good idea to send people to live on Mars?
5. What sort of things do you have to do to look after a dog?
6. Why do people go to Scotland on holiday?

In the study with healthy participants, a written prompt was presented on a computer screen prior to each speech elicitation period. The prompt was removed from the screen when the participant was ready to begin speaking and they were allowed to speak for 60s. When testing patients, however, the prompts were read aloud by the researcher and no time limit was imposed on responses. We did not use a time limit because we expected speech rate to be reduced in our patients and therefore that they would produce significantly less speech than controls if only allowed to speak for 60s. Instead, patients were encouraged to speak for as long as chose to about each subject. If they provided little information spontaneously, the researcher encouraged them to elaborate with a neutral comment like “Can you say anything else about that?”. To ensure that potential differences between groups in quantity of speech could not account for our results, we recorded the length of each response in number of words and included this as a covariate in all analyses (see Results).

Our main analyses used speech from the full period that participants spoke for. Because patients were allowed to speak for longer than controls, we also performed supplementary analyses in which we computed coherence values using only speech produced in the first 60s of each patient response (thus matching the time period to the control group). The results were very similar to those we report below (see Supplementary Materials for details), indicating that observed effects were not due to differences in the time allowed to respond.

### Speech processing and computation of coherence

Spoken responses were digitally recorded for later transcription. Non-lexical fillers (umm, ah etc.) were not transcribed and pauses were not marked. All lexical items were transcribed but some editing of transcripts was performed prior to further analysis. Specifically, following Glosser & Deser (1990), we removed false starts and aborted utterances (“We *wanted to*…we always went to the shops”; removed speech shown in italics), immediate perseverations of individual words or phrases (“But it’s not, *it’s not* got air there”), and comments about the task itself or the patients’ language problems (“*I don’t know what the word is*”). We did this because we wanted to take a functional approach that analysed the coherence of the overall message conveyed by patients, while minimising the effect of lexical retrieval problems or other microlinguistic deficits. 9% of words produced by SA patients were removed as a result of this process. The same editing was applied to controls’ speech, with 1.6% of their total word count removed.

Our main dependent variables were computed measures of global and local coherence (GC and LC), as described below. Other markers of lexical-semantic content were also computed and were included in supplementary analyses (see Statistical Analyses). Measures of GC and LC were generated using an automated computational linguistic approach, first described by Hoffman et al. (2018). Analyses were implemented in R; the code is publicly available and can easily be applied to new samples (https://osf.io/8atfn/). Our approach made use of latent semantic analysis (LSA) (Landauer & Dumais, 1997). LSA provides the user with vector-based representations of the meanings of words, which can be combined linearly to represent the meanings of whole passages of speech or text (Foltz et al., 1998). Using similar methods to other researchers (Elvevag et al., 2007; Foltz, 2007), we used these representations to characterise the content of each speech sample and to quantify its coherence. Coherence calculated in this way has high internal reliability and test-retest reliability and is highly correlated with human ratings of coherence (Hoffman, Loginova, et al., 2018).

The computation process is illustrated in Supplementary Figure 3. Our overall strategy was to divide each speech sample into smaller windows (of 20 words each) and to use LSA to generate vector representations of the semantic content of each window. Coherence was then assessed with a moving window approach. LC was assessed by measuring the similarity of the vector for each window with that of the patient’s previous window. Therefore, in common with other researchers (Elvevag et al., 2007; Foltz, 2007), we define LC as the degree to which adjoining utterances convey semantically related content. A low LC value would be obtained if a patient switched abruptly between topics during their response. We used the cosine between vectors as the measure of their similarity, so a value of 0 indicates no semantic relationship between windows and 1 indicates identical content.

GC was assessed by comparing the semantic content of each window with a vector representing the prototypical semantics produced by healthy participants to the same prompt (GC). To generate this prototypical representation, we took all the responses made by control participants and computed an LSA representation for each one. We then averaged these to give a composite vector that represented the typical semantic content that healthy individuals produced when responding to the prompt^2^ (for further details of the LSA space and averaging procedure, see Hoffman, Loginova, et al., 2018). For example, the composite vector for the prompt “Describe the steps you would need to take if going somewhere by train” would be similar to the vectors for *train, railway, ticket, station* and so on, as these words were frequently used in responses to this prompt. The GC for each window was defined as the similarity between its vector and the composite vector. Therefore, GC was a measure of how much a patient’s response matched the typical semantic content of responses to that prompt. A low GC value would be obtained if a patient tended to talk about other topics that were semantically unrelated to the topic being probed. Thus, our measure of GC captured the degree to which patients maintained their focus on the topic under discussion, in line with the definition used by other researchers (Glosser & Deser, 1992; Wright et al., 2014).

Once GC and LC had been calculated, the window moved one word to the right and the process was repeated, until all windows had been assessed. GC and LC values were averaged across windows to give overall values for each response. It is important to note that, although GC and LC are both measured on a scale of 0 to 1, their values are not directly comparable because of differences in the way in which they are calculated. GC values tend to be higher than LC values because they are the result of comparisons to composites derived from many hundreds of words.

### Statistical analyses

Group analyses compared performance in the SA group (not including the non-aphasic RH patient) with the age-matched healthy control group. At the group level, analyses were performed using linear mixed effects (LME) models. Accuracy on the semantic control task was analysed using a generalised LME model with a logit link function. The model had a 2 × 2 × 2 (group × task × control demands) factorial structure and included random intercepts for participants and test items, as well as all random slopes for all factors varying within-unit (Barr et al., 2013). The significance of fixed effects was assessed using a likelihood ratio test to compare the full model with a reduced model identical except for the removal of the effect of interest (Barr et al., 2013).

To test for group differences in coherence, LME models were estimated separately for GC and LC values, with a factorial manipulation of group (SA vs. controls, again excluding the RH patient) and including response length as a control predictor (since longer responses may be more likely to deviate off-topic; Hoffman, Loginova, et al., 2018). These models included random intercepts for participants and prompts and random by-probes slopes for the effect of group. Effects of group were assessed using a t-test with Satterthwaite approximation of degrees of freedom, as there were a smaller number of observations here which could cause likelihood ratio tests to be anti-conservative (Huber et al., 2015). To determine whether the coherence scores of individual patients differed from the control group, we repeated the LME analyses, each time comparing the control group to a single patient. This method extends the modified t-test approach to the LME framework allowing us to take item effects into account when testing for impairments (Huber et al., 2015). We also computed correlations between coherence measures and semantic task performance in the full patient group.

Finally, we calculated seven other markers of the lexical-semantic properties of speech: the mean frequency, concreteness, age of acquisition, semantic diversity and length of words produced, type:token ratio and the proportion of closed-class words (see Supplementary Materials for full details). To analyse these data, we performed a principal components analysis to reduce the individual measures to four latent factors (Following the same method as our previous studies; Hoffman, 2019; Hoffman, Loginova, et al., 2018). Consistent with previous work, the four factors appeared to index specificity of semantic content, complexity of vocabulary, coherence and lexical content (see Table 4 for factor loadings). We performed LME analyses to the scores on each factor to determine whether patients differed from controls on each factor, and tested whether correlations with semantic performance were present.

### Data availability

The data reported in this paper are available at https://osf.io/cvuqz/

## Results

### Background neuropsychological tests

Scores on the background neuropsychological tests are presented in Table 2. In all tables and figures, SA patients are ordered by the severity of their semantic control impairment (i.e., mean performance on the high control conditions of the semantic control task, as reported below). SA patients showed evidence of multimodal semantic impairments on background testing, in that each patient performed below the normal range in at least one verbal and one non-verbal semantic task. Semantic impairment in SA is typically accompanied by domain-general executive impairments (Jefferies & Lambon Ralph, 2006; Thompson et al., 2018) and this was also the case here. Patients showed impairment on tests of general executive function, such as the Brixton test of spatial anticipation, the Trails test part B and Raven’s progressive matrices. All SA patients performed below the normal range on at least one of these tests, with the exception of SA1 and SA2 (who also showed the most intact performance on tests of semantic control; see below). The right-hemisphere patient, RH1, performed well on the semantic tasks, displaying mild impairment only on the synonym judgement task.

**Table 2:** Scores on background neuropsychological tests

|  | Normative mean (cut-off) | RHI | SA1 | SA2 | SA3 | SA4 | SA5 | SA6 | SA7 |
| --- | --- | --- | --- | --- | --- | --- | --- | --- | --- |
| Cookie theft (words per minute) |  | NT | 38 | 58 | 54 | 60 | 77 | 41 | 97 |
| Category fluency (8) | 115.1 (76) | 18<sup>a</sup> | 69* | NT | 80 | 14* | NT | 46* | 24* |
| Letter fluency (F,A,S) | 41.1 (19) | 17<sup>b</sup> | 12* | NT | 16* | 3* | NT | 18* | 19* |
| *Semantic Cognition* |  |  |  |  |  |  |  |  |  |
| Synonym judgement /96 | 94.5 (91) | 88* | 81* | 87* | 78* | 66* | 64* | 81* | 81* |
| Camel & Cactus (pictures) /64 | 59.1 (51) | 56 | 61 | 60 | 53 | 45* | 49* | 58 | 44* |
| Canonical Object use /37 | - | - | 37 | 36 | 37 | 37 | 32 | 25 | 30 |
| Non-canonical object use /37 | 36.0 (34) | - | 32* | 32* | 26* | 34 | 17* | 12* | 12* |
| Picture naming /64 | 62.3 (59) | 64 | 46* | 14/16* | 56* | 60 | NT | 13/16* | 21* |
| Word-picture matching /64 | 63.8 (62) | 64 | 63 | 63 | 64 | 62 | 11/16* | 15/16* | 46* |
| *Cognition and Executive* |  |  |  |  |  |  |  |  |  |
| Brixton test of spatial anticipation /54 | 39 (30) | 21* | 39 | 45 | 31 | 24* | 27* | 41 | 29* |
| Trail-making test Part A /24 | 23.9 (23) | 24 | 24 | 24 | 24 | 24 | 23 | 24 | 23 |
| Trail-making test Part B /23 | 23.5 (21) | 22 | 21 | 23 | 19* | 1* | 16* | 14* | 7* |
| Raven's coloured progressive matrices /36 | 29.7 (20) | 30 | 33 | 29 | 31 | 19* | 30 | 36 | 29 |
| Forward digit span | 6.8 (4) | 7 | 6 | 3* | 5 | 6 | 2* | 3* | 8 |
| Backward digit span | 4.8 (3) | 4 | 4 | NT | 3 | 2* | 1* | 3 | 4 |
| VOSP dot counting /10 | 9.9 (9) | 10 | 10 | NT | 10 | 10 | NT | 10 | 8* |
| VOSP number location /10 | 9.4 (7) | 10 | 8 | NT | 5* | 10 | NT | 10 | 8 |

Control data were obtained from published norms. Minimum control score indicates cut-off below which performance is considered abnormal (two standard deviations below the mean if no other threshold was provided). Patient scores below this level are indicated by \*. For some of the semantic tests, shortened 16-item versions were administered to some patients. <sup>a</sup> = one category "animal", <sup>b</sup> = one letter "S". NT = not tested. VOSP = Visual Object and Space Perception battery.

### Tests of semantic control

Semantic control was assessed using a forced-choice semantic judgement task with a 2 × 2 design that manipulated the type of semantic judgement (global vs. specific feature) and the need for controlled semantic processing. Accuracy on this task is presented in Figure 1. The data were analysed with generalised mixed effects models. SA patients were significantly impaired on the task as a whole, compared with the healthy control group (*B* = 0.58, *se* = 0.22, *p* = 0.004). There was a main effect of the semantic control manipulation (*B* = −0.95, *se* = 0.20, *p* < 0.001), indicating poorer performance in the high control conditions. There was also a marginal interaction between control demands and group (*B* = -0.39, *se* = 0.19, *p* = 0.062), suggesting that manipulating the need for semantic control may have had a larger effect on the patients than on the controls. There was no effect of task and no interactions with other factors, suggesting that the two different manipulations of control in this experiment (controlled retrieval vs. semantic selection) had similar effects on our participants. These data establish that patients in the SA group had difficulty in single-word semantic processing when semantic control demands were high, to varying degrees of severity. In contrast, RH1 performed within the normal range on these tasks.

**Figure 1:**
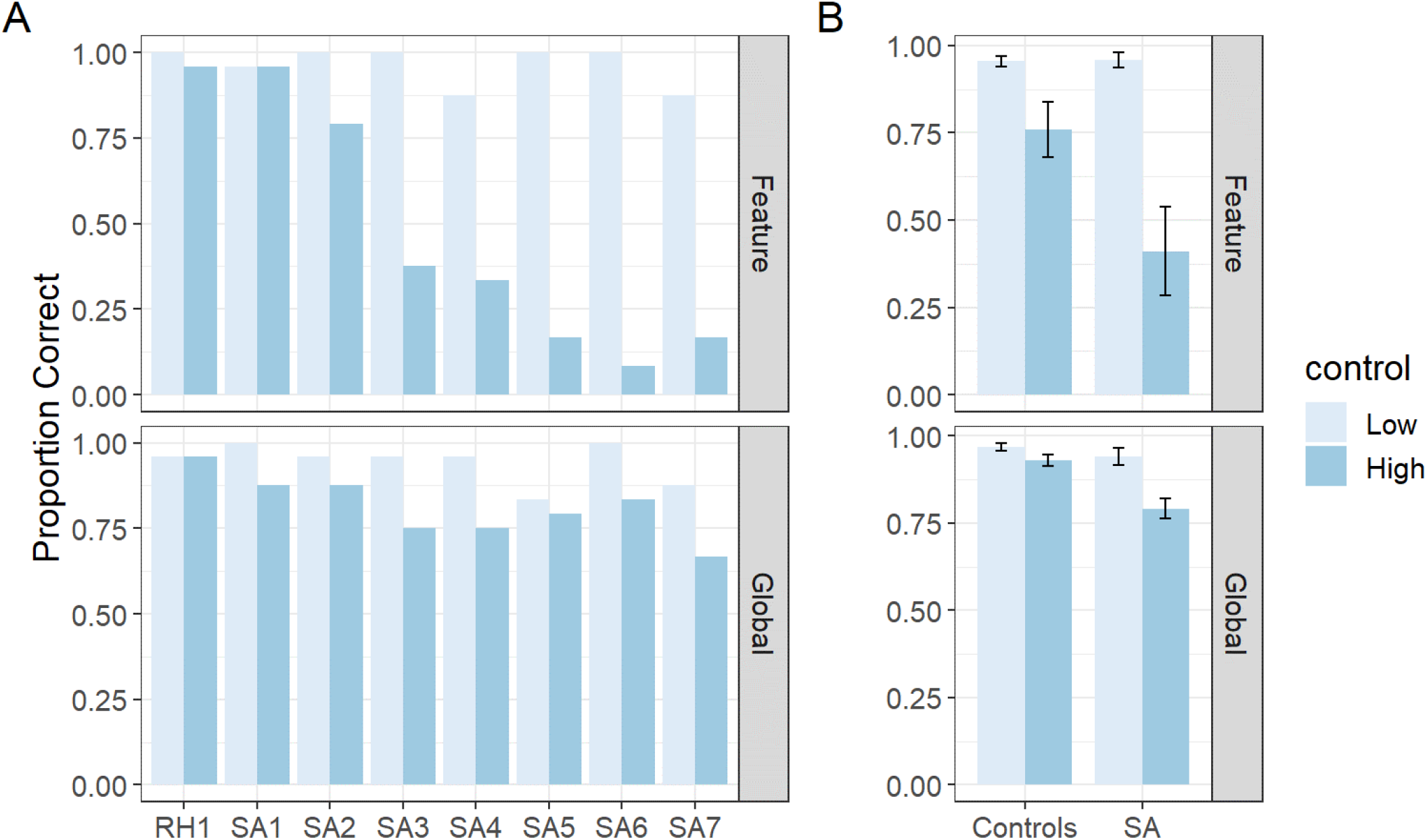
Performance on semantic control tasks. (A) Performance in each patient, (B) means for SA patients and controls. Bars indicate one standard error of the mean. SA = semantic aphasia.

### Speech elicitation

Basic information on the speech samples collected from patients is provided in Table 3. We presented either five or six prompts to each patient, depending on the time available for testing. For SA3, we were only able to analyse data from four prompts for analysis because in the other two cases she failed to produce enough words to provide reliable coherence data (7 and 17 words only). The mean duration of patients’ responses varied between 62 and 115s (in contrast, a 60s time limit was imposed on controls). Despite this, SA patients produced fewer words on average than controls (*t*(17) = 3.09, *p* = 0.007) because they spoke at a much slower rate (*t*(17) = 4.89, *p* < 0.001). In contrast, RH1 was above the control mean for response length and speech rate.

**Table 3:**
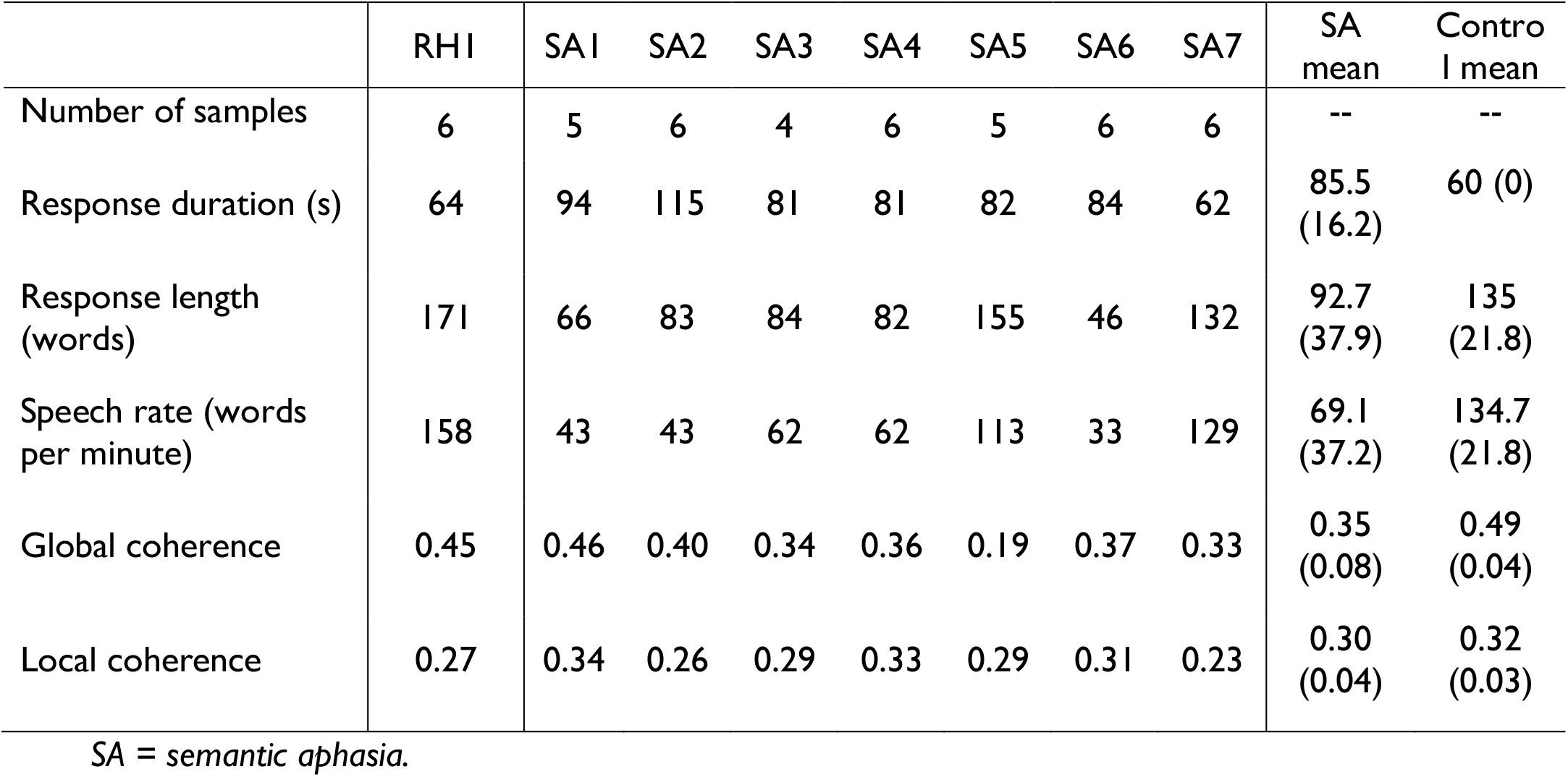
Characteristics of speech samples

Examples of SA patients’ responses are given are provided in Supplementary Materials. Patients appeared to frequently deviate from the topic about which they were asked, behaviour which we analyse formally in the next section.

### Speech coherence

Our formal assessment of coherence involved using previously-validated computational linguistics methods to quantify the global coherence (GC) and local coherence of each response (LC). Mean GC and LC for each patient is shown in Table 3, with SA and Control means in Figure 2. LME analyses of these data (including length of responses as a covariate) revealed that SA patients were significantly less globally coherent than controls (*t*(11.4) = 5.11, *p* < 0.001) but not less locally coherent (*t*(24.4) = 1.45, *p* = 0.16). At the level of individual patients, all patients except SA1 and SA2 showed significantly impaired GC scores relative to controls (*p* < 0.05). SA7 was the only patient to exhibit an LC score that was significantly lower than the control group (*p* < 0.05). RH1 was not impaired on either coherence measure (*p* > 0.05).

**Figure 2:**
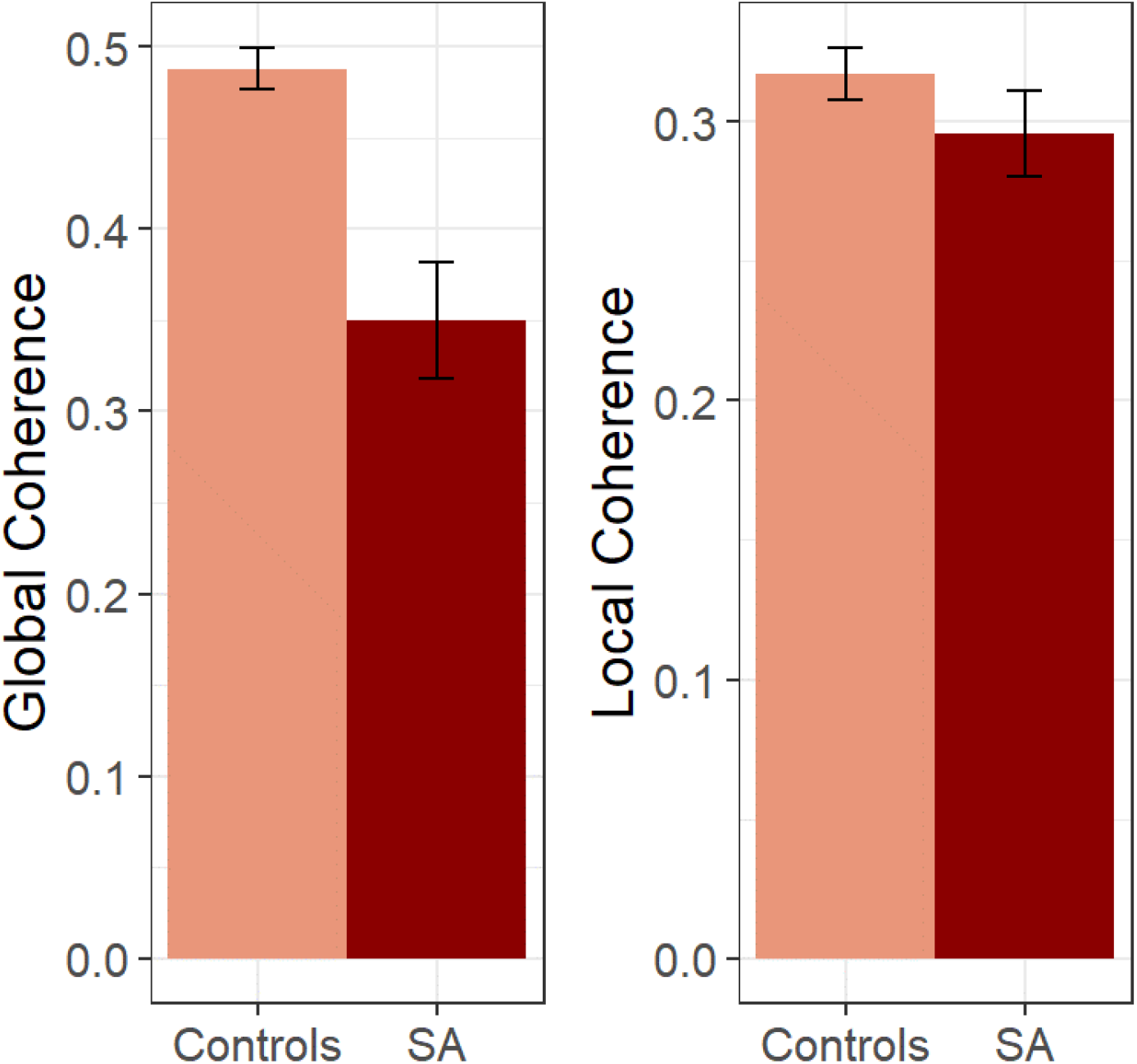
Mean coherence levels for SA patients and controls. Bars indicate one standard error of the mean. SA = semantic aphasia. Note that, due to differences in the calculation method, global and local coherence values are not measured on equivalent scales and cannot be compared directly.

Correlations between the coherence measures, response length and scores on the high-control semantic conditions are shown in Figure 3. There was a strong tendency for patients who performed better on the high-control semantic tasks to score more highly for GC in speech (Incongruent Features: *r* = 0.75, *p* = 0.03; Weak Associations: *r* = 0.56, *p* = 0.14). In contrast, semantic control performance did not predict LC values, nor did it predict the average length of responses.

**Figure 3:**
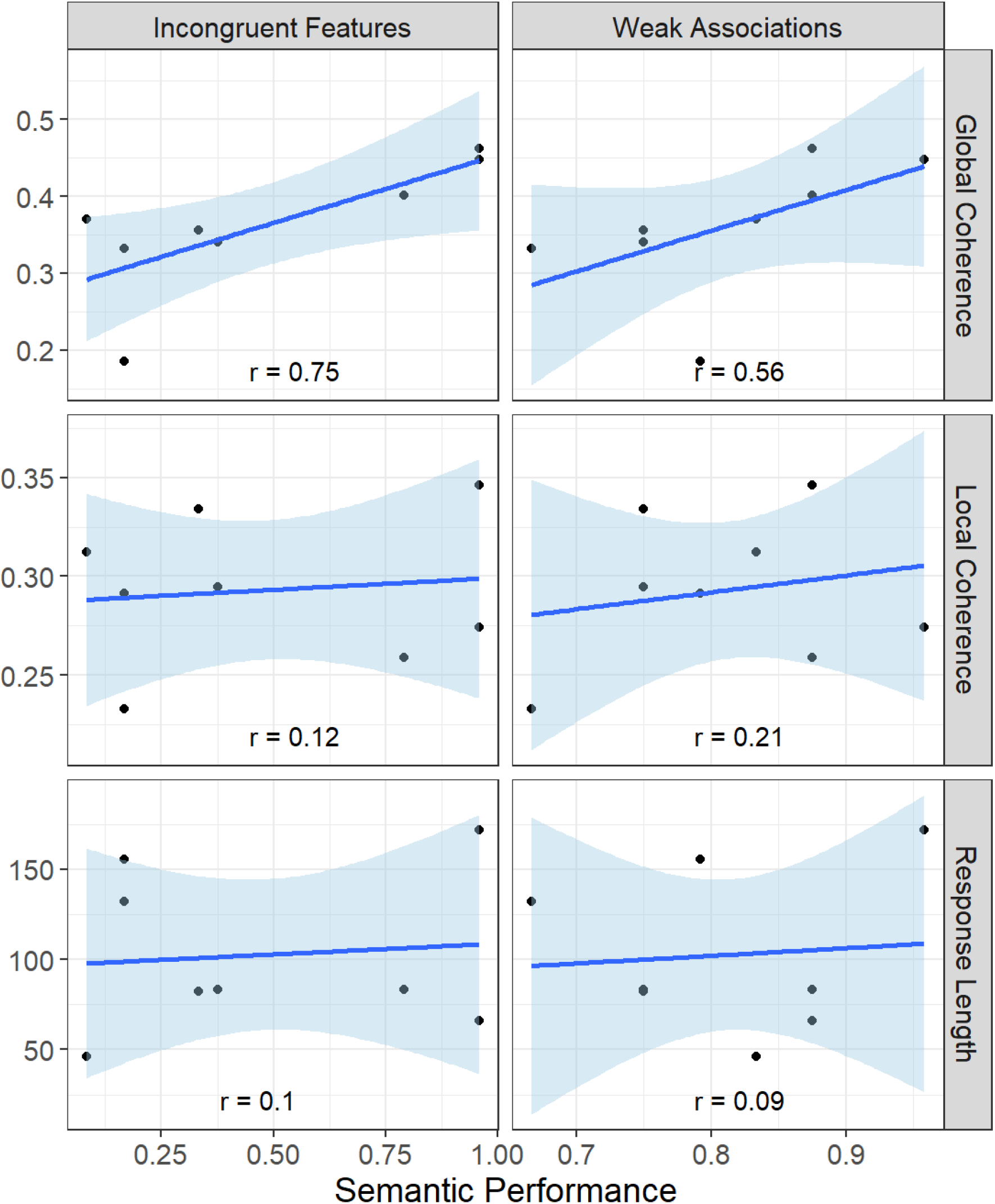
Relationships between coherence, response length and semantic control performance in the patients

### Other characteristics of speech

Finally, we computed a range of other lexical-semantic speech markers to determine the degree to which semantic control impairment affected other aspects of speech content (see Supplementary Materials for details). Following the same method as previous studies (Hoffman, 2019; Hoffman, Loginova, et al., 2018), we performed a principal components analysis on these data, which identified four factors corresponding to distinct aspects of speech production (see Table 4 for factor loadings):

1. Semantic specificity, the tendency to produce highly concrete, low frequency words with low semantic diversity
2. Complexity of vocabulary, the degree to which participants produced long, low frequency, late-acquired words
3. Coherence, which was strongly related to the GC and LC measures analysed above
4. Lexical content, the degree to which participants produced a large proportion of open-class content words.

**Table 4:** Factor loadings in principal components analysis of speech characteristics, and mean factor scores in each group? Loadings with absolute values >0.4 are shown in bold. SA = semantic aphasia.

| Measure | Factor 1:<br>Semantic<br>specificity | Factor 2:<br>Vocabulary | Factor 3:<br>Coherence | Factor 4:<br>Lexical<br>content |
| --- | --- | --- | --- | --- |
| Noun frequency | **-.61** | **-.49** | -.08 | -.07 |
| Noun semantic diversity | **-.88** | -.15 | .05 | -.09 |
| Noun concreteness | **.72** | -.28 | .29 | .03 |
| Type:token ratio | **-.59** | .29 | .39 | .19 |
| Noun age of acquisition | .01 | **.91** | -.13 | -.23 |
| Noun phonemic length | .05 | **.82** | -.00 | .13 |
| Global coherence | -.22 | .14 | **.77** | -.10 |
| Local coherence | .02 | -.21 | **.87** | -.06 |
| % closed class | .09 | .09 | .11 | **-.99** |
| *Mean factor scores by group:* |  |  |  |  |
| Controls mean (s.d.) | -0.01 (0.30) | 0.45 (0.61) | 0.49 (0.27) | 0.29 (0.50) |
| SA mean (s.d.) | 0.04 (1.14) | -0.49 (0.61) | -0.46 (0.58) | -0.21 (0.78) |
| Effect of group | $t(12.4) = 1.35,$<br>$p = 0.202$ | $t(14.1) = 3.78,$<br>$p = 0.002$ | $t(11.8) = 3.94,$<br>$p = 0.002$ | $t(16.7) = 1.94,$<br>$p = 0.069$ |

Table 4 also shows the mean factor scores for each group and the effects of group, assessed using LME models of the same form used to analyse GC and LC. In addition to differing from controls on the latent coherence factor, SA patients also tended to use less complex vocabulary than controls. Correlations between patients’ performance on tests of semantic control and their scores on the latent speech factors are shown in Figure 4. The coherence factor showed the strongest correlations with semantic control ability, replicating the main analysis of GC. However, there was also a weaker tendency for patients with more semantic control ability to use more complex vocabulary. This suggests that semantic control ability influences the coherence of speech more than it does other aspects of speech content (in line with Hoffman et al.’s (2018a) findings in healthy participants).

**Figure 4:**
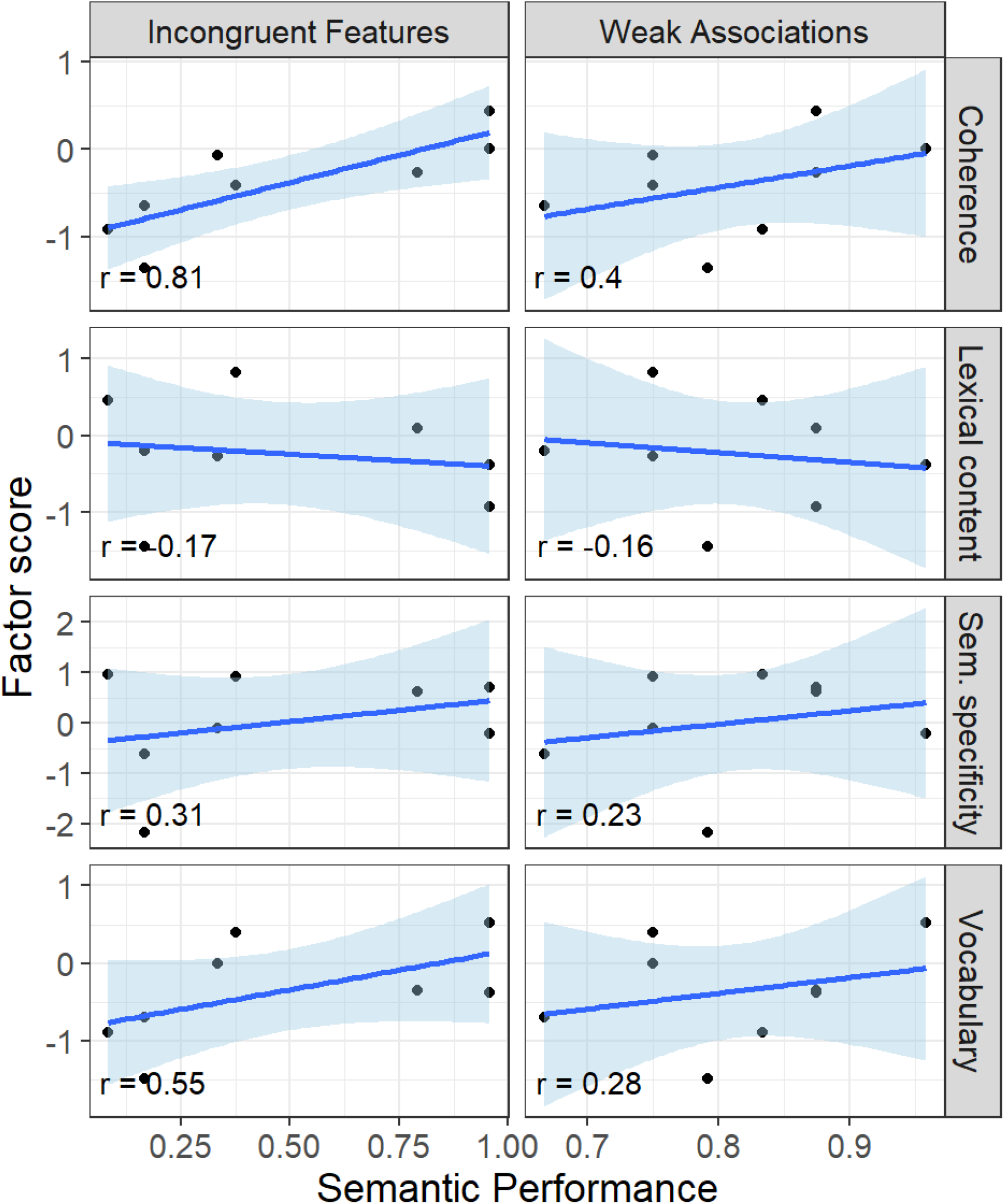
Relationships between semantic control performance and latent speech factors in the patients

## Discussion

Maintaining coherence when speaking is a critical skill which may depend on the ability to retrieve and select currently-relevant knowledge from semantic memory (known as semantic control). We investigated the coherence of connected speech in a case-series of stroke patients with multimodal semantic impairments. One previous study found associations between multimodal semantic deficits and global coherence in Alzheimer’s disease (Laine et al., 1998). Our stroke patients also showed gross impairments to global coherence: compared to controls, their spoken responses were less relevant to the topics we asked them to speak about. Importantly, however, these deficits were strongly correlated with performance on tests of semantic control, with the patients with the poorest semantic control exhibiting the lowest global coherence values. Other aspects of speech production were also impaired but none showed such a strong correlation with semantic control abilities. Our findings provide converging support for the idea that semantic control processes are central to maintaining global coherence in speech and indicate that poorly coherent speech is potentially a major issue for patients with deficits of semantic control.

Before discussing our preferred interpretation for these results, we consider the alternative possibility that observed deficits in global coherence were a side-effect of microlinguistic impairments in our patients. Language impairments at the microlinguistic level are sometimes associated with poor coherence (Andreetta et al., 2012; Andreetta & Marini, 2015; Christiansen, 1995; Wright & Capilouto, 2012). However, such deficits are unlikely to provide a full explanation for the impaired coherence observed here, for a number of reasons. First, because we were interested in assessing the content of speech at the functional, macrolinguistic level, prior to analysis we removed elements of speech that might occur more frequently as a consequence of aphasic deficits (e.g., aborted utterances, perseverative speech). This step minimised the effect of interruptions to fluent speech by eliminating them from coherence computations. Second, we found that while performance on tests of semantic control predicted levels of global coherence, these tests were less strongly correlated with other lexical-semantic characteristics of speech content, including response length, complexity of vocabulary and semantic specificity. Thus our data suggest that, although the speech of SA patients differs from that of healthy people in a number of ways, it is the impairment in coherence that most strongly related to poor semantic control.

The final point concerns what type of disruption to global coherence one would expect as a consequence of microlinguistic deficits. In two of the largest studies of coherence in aphasia (Andreetta et al., 2012; Andreetta & Marini, 2015), four types of utterance were coded as errors of global coherence: tangential utterances, conceptually incongruent utterances, propositional repetitions and filler utterances (e.g., “yes, I get it”). Of these, only repetitions and fillers occurred at an elevated rate in aphasic patients. This suggests that the presence of aphasia causes speech to become more repetitive and less informative, but does not in itself cause patients to provide tangential or off-topic information (Andreetta & Marini, 2015). In contrast, our patients showed global coherence deficits because they provided information that was semantically distant from the original topic, and not because they were repetitive or uninformative. However, within the methods we have used, the presence of repetitions and filler utterances has little impact on global coherence.^3^

Our patients exhibited significant impairments to global coherence but not to local coherence, when compared with healthy controls. In other words, they were able to maintain semantic relationships between consecutive utterances, but they were poor at ensuring those utterances remained relevant to the topic at hand. The result was that their verbal output often had the character of an undirected stream of consciousness: they began producing topic-relevant information but over time, drifted away from the topic they had been asked about (see Supplementary Materials for examples). Our interpretation of this behaviour is that *automatic* semantic retrieval processes are relatively intact in these cases. When speaking, they successfully activate a chain of meaningful associations which drive their speech output. However, in order to maintain focus on the topic at hand, top-down control processes must act on the information retrieved from semantic memory, to ensure that information selected for production is relevant to the topic under discussion. Disruption to these selection processes appear to be responsible for the patients’ global coherence deficits.

A distinction has often been drawn between two distinct forms of semantic control: *semantic selection* of task-relevant knowledge vs. *controlled retrieval* of less salient knowledge (Badre & Wagner, 2007). Our account of the SA patients’ coherence deficits holds that the selection element plays the key role in maintaining global coherence. This is consistent with our previous study in healthy participants (Hoffman, Loginova, et al., 2018) and with an fMRI study, again in healthy older people, indicating that high coherence during speech is correlated with increased activation in the pars triangularis (BA45) region of left inferior frontal gyrus (Hoffman, 2019). This area is strongly implicated in semantic selection (Badre et al., 2005; Badre & Wagner, 2007; Thompson-Schill et al., 1997). In the present study, however, we found that tasks probing selection and controlled retrieval showed similar correlations with global coherence. SA patients typically show parallel impairments to both of these aspects of semantic control, making it difficult to tease apart contributions of each in this patient group. However, the converging evidence across these studies suggests that poor selection of semantic information may be critical in understanding coherence impairments.

Another outstanding question is the degree to which the semantic selection processes we measured here overlap with executive functions in other cognitive domains. This is not an issue we were able to address directly in this study because patients did not complete comprehensive assessments of executive function in other domains. However, other studies in SA patients indicate that general executive deficits typically co-occur with, and are correlated with, semantic control impairments (Jefferies & Lambon Ralph, 2006). Furthermore, patients who present with a general dysexecutive syndrome also show the hallmarks of a semantic control disorder (Thompson et al., 2018). It is not clear the degree to which these co-occurring deficits indicate a common functional system, or are simply a consequence of concomitant damage to distinct but neurally proximate systems. However, recent work in healthy individuals suggests that semantic selection ability patterns closely with performance on non-semantic executive tasks (Hoffman, 2018). Thus, it is possible that inhibition of irrelevant semantic information is closely related to inhibitory functions in other cognitive domains. This would be consistent with the general view that domain-general executive functions support the regulation of speech at the macrolinguistic level (Alexander, 2006; Barker et al., 2020; Barker et al., 2017; Kintz et al., 2016).

Our account of deficits in semantic selection has much in common with the concept of an “idea selection” deficit, which has been proposed to explain language impairments in dynamic aphasia (Barker et al., 2020; Robinson et al., 1998; Robinson et al., 2010). Dynamic aphasia is a rare condition in which spontaneous production of propositional speech is severely reduced, even though patients have intact language processing at the microlinguistic level. Patients with this condition are thought to have difficulty selecting a single proposition for production when multiple ideas are strongly activated. Empirical evidence for this proposal has principally come from sentence-level production tasks in which a number of potential, equally valid, propositions compete for selection (e.g., completion of sentences with low cloze probabilities, like “there was a nothing with the…”). Patients with dynamic aphasia frequently fail to produce any response under these circumstances, despite performing well in conditions with greater constraint (“When you go to bed, turn off the.”). Although the theoretical account put forward for dynamic aphasia has much in common with our account of coherence deficits in SA, there also appears to be some differences in the observed deficits. Unlike cases of dynamic aphasia, our SA patients readily produced ideas in response to topic prompts (although they did produce fewer words than controls in some cases). They failed, however, to ensure that the information they produced was topic-relevant. This suggests that, rather than an inability to select per se, SA patients failed to ensure that the selection process was guided by current context and goals. Further investigation of the relationship between dynamic aphasia and semantic control impairments may be helpful in understanding the cognitive mechanisms that support selection of semantic information for language production.

For the present study, we deliberately used a relatively unconstrained speech elicitation task. Patients were given a topic to speak about and were required to structure their own response with minimal support from the experimenter. We chose this method to provide a strong test of patients’ ability to regulate their own speech output. However, it is worth noting that SA patients show positive effects of increasing task constraints across a range of semantically-driven tasks and we would expect to see similar effects here (Corbett et al., 2011; Noonan et al., 2010). For example, in everyday life the patient’s conversational partners may play an important role in providing verbal and non-verbal cues that direct them back towards topic-relevant discourse. There is a parallel here with studies in healthy older adults, in which coherence impairments are greatest when verbal topic prompts are used and less prominent when participants are asked to describe pictures or comic strips, where there is a visual reminder of the subject matter throughout (James et al., 1998; Wright et al., 2014). One way to improve coherence in this patient group may therefore be to provide external cues that maintain their focus on the current topic.

In conclusion, the unique contribution of this research is to show the nature of impairments to propositional speech brought by semantic control impairments. SA patients have particular difficulties maintaining global coherence, such that their unregulated retrieval of semantic information leads them “off the rails” and progressively away from the topic at hand. Better understanding of these processes will be critical in uncovering the root causes of conversational difficulties experienced by aphasic patients and will be useful in guiding rehabilitation in hospital, clinics and in the home.

## Data Availability

The data reported in this paper are available at https://osf.io/cvuqz/

## Acknowledgements

We are grateful to the patients who generously offered their time to take part in this study.

## Footnotes

1 Two other SA patients attempted to take part in the study but were unable to complete the speech elicitation task. Their speech was considerably less fluent than the patients who successfully completed the task (on the Cookie Theft description, they produced 9 and 12 words per minute, cf. a mean of 61 words per minute in the patients who completed the task).

2 However, when using this procedure to measure GC of a control participant, we generated a new composite that excluded that participant’s response. This was important to ensure that the speech used to generate the composite representation was independent of the speech currently being analysed.

3 A repetitive response will not generate a low coherence score so long as the repeated information is semantically relevant to the topic being discussed. As for filler utterances, they tend to convey little semantic content thus have little effect on the computation of coherence. This is because when the semantic content of speech passage is calculated, the contribution of each word is weighted by its entropy in the British National Corpus (Hoffman et al., 2018). Words that are semantically uninformative (e.g., the, yeah) therefore receive low weightings when computing the semantics of the passage.

